# Prevalence of Returnable Genetic Results Based on Recognizable Phenotypes among Children with Autism Spectrum Disorder

**DOI:** 10.1101/2021.05.28.21257736

**Authors:** Somer L. Bishop, Audrey Thurm, Elise Robinson, Stephan J Sanders

**Author notes:** **Please address correspondence to**: Stephan J. Sanders, University of California, San Francisco, 1550 Fourth St, 445 Box 2822, San Francisco, CA 94158.

## Abstract

The importance of extensive genetic testing of autism spectrum disorder (ASD) cases has been demonstrated repeatedly in research settings but such testing in clinical settings remains sporadic. Determining which individuals should be prioritized for expensive tests remains a challenge. Several guidelines have been released relating to clinical genetic evaluations and testing in the context of ASD and these guidelines may be informed by the results of genetic testing in large research cohorts. The current study summarizes findings from over 2,000 individuals with ASD who received genetic testing, including microarray and exome testing, through the Simons Simplex Consortium. A returnable genetic result is identified in 10% of cases, however, this yield increases based on four readily accessible phenotypes: female sex and the presence of intellectual disability, seizures or delayed walking. Combinations of these factors increase return rate further, with some combinations yielding a return rate over 50%. In conclusion, these four phenotypes provide a simple approach to prioritize genetic testing in a clinical setting and inform future clinical guidelines. Providing a systematic approach to decisions about who to test removes barriers for, and therefore decrease disparities in, reimbursable genetic testing for individuals diagnosed with ASD.

## Introduction

A substantial proportion of children with Autism Spectrum Disorder (ASD) have an identifiable genetic condition that causes or substantially contributes to their symptoms. Once thought to be extremely rare, research cohorts have identified etiologically significant genetic testing results in at least 10% of individuals with ASD.^1^ However, despite the high diagnostic yield, the use of genetic testing as part of clinical evaluation of patients with ASD is inconsistent, even in the presence of phenotypes associated with genetic disorders in ASD (e.g., motor delay, seizures, intellectual disability, female sex).^2-4^ Moreover, insurance companies often do not cover the cost, and out of pocket expenses for testing can be exorbitant. This not only decreases the likelihood that families will pursue testing, but also may make clinicians hesitant to encourage it.

To help prioritize clinical decisions about genetic testing in individuals with ASD, we undertook an analysis of the Simons Simplex Collection (SSC),^2^ the largest ASD research cohort with consistent genomic and phenotypic data, to estimate the percent of cases with returnable genetic results (i.e. those reported to a clinician/patient) based on key recognizable phenotypes.

## Methods

We identified 2,218 SSC probands for whom exome sequencing and microarray data were available, along with phenotypic data on four previously identified predictors of genetic disorders in ASD: sex, intellectual disability (IQ ≤ 70), delayed walking (≥ 18 months), and presence of afebrile seizures.

A returnable genetic result was defined as (i) a *de novo* copy number variant or a *de novo* variant in an ASD-associated or developmental-delay-associated gene leading to protein truncation (n genes= 328) or (ii) damaging missense variant (missense badness, PolyPhen-2, constraint [MPC] score ≥ 2) in that same set of genes.^1,2,5^ Ranking the phenotypic markers from the most (delayed walking) to least (intellectual disability) discriminating, we assessed the proportion of cases with a returnable genetic result in relation to each phenotypic marker independently and in combination.

## Results

A returnable genetic result was identified in 234 out of 2,218 (10.6%) probands, including 87 copy number variants, 92 protein-truncating variants, and 57 missense variants (236 total, two probands had multiple returnable results, Table S1). Only 72 of the 1,220 (5.9%) males without walking delay, seizures, or intellectual disability had a returnable genetic result. The presence of one or more of the four phenotypic markers increased the returnable yield to 16.2% (162 out of 998 cases), which further rose to 60% (15 out of 25 cases) in the presence of delayed walking and seizures (Fig. 1).

**Figure 1.**
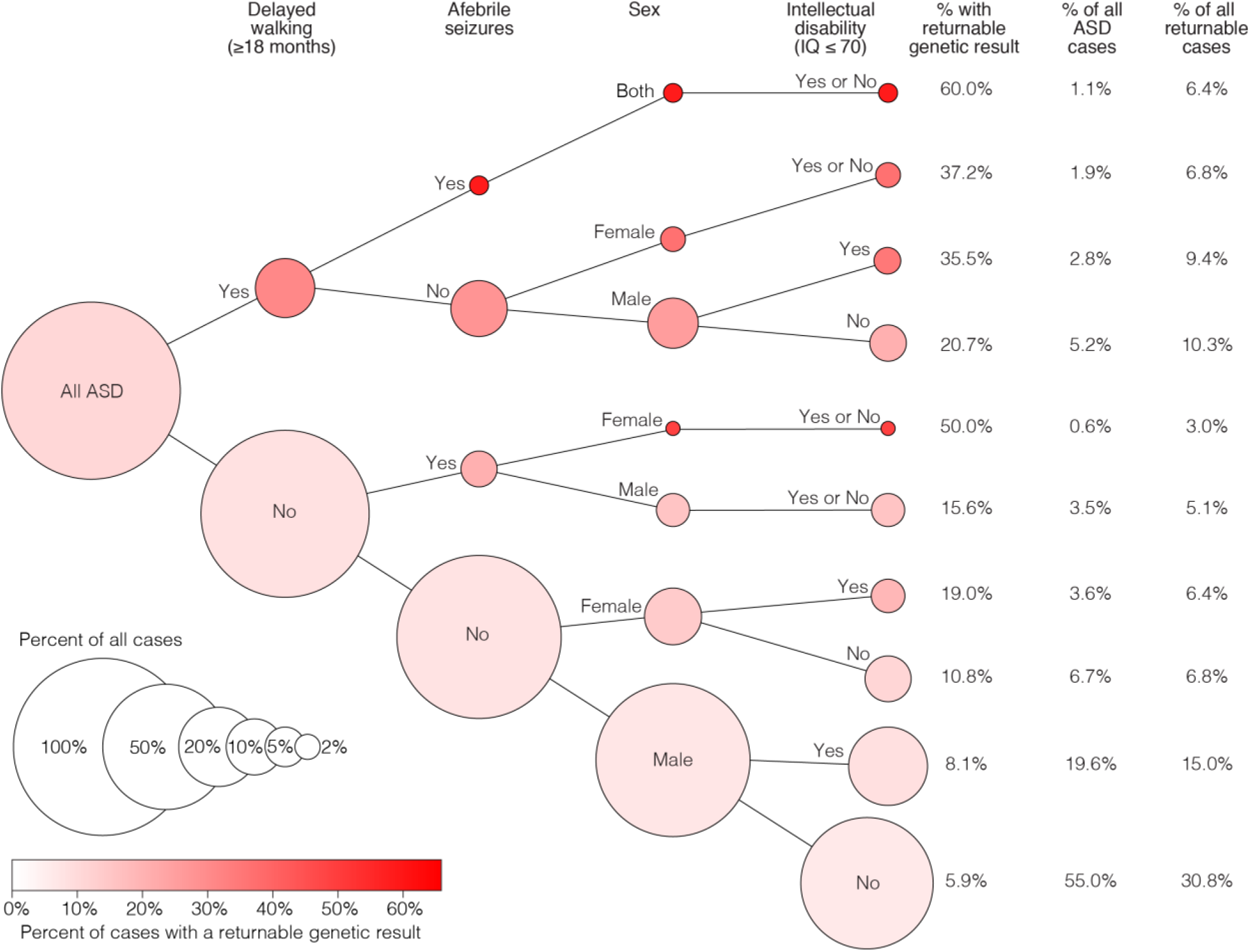
Flowchart of the yield of returnable genetic results of exome and clinical microarray genetic testing in ASD based on four key phenotypic markers. Outcomes are not split by intellectual disability phenotype (‘Yes or No’) if there are fewer than 50 individuals or less than 1% difference in the percentage with a returnable result.

## Discussion

Information about genetic etiology is important for families. Genetic conditions convey putative risk for preventable and/or treatable conditions, provide support through dedicated family groups, and enable enrollment in clinical trials, some of which are already underway to test gene-specific therapeutic agents (e.g. https://clinicaltrials.gov/ct2/show/NCT04259281). Genetic results are also critical for scientific progress, including studies of natural history and long-term outcomes, comorbidities, and response to therapies.

Clinical microarray and exome sequencing have been recommended after a diagnosis of ASD, but are not routinely ordered by clinicians or covered by insurance. Results of this analysis indicate that, among patients with ASD, returnable genetic results are found in 1 in 5 females, and more than 1 in 4 males and females presenting with a history of delayed walking. While a clinical genetics evaluation is recommended on the basis of ASD diagnosis alone,^6^ information about easily ascertainable phenotypes (i.e., age of walking, presence of seizures and/or intellectual disability, and biological sex) may provide even further justification to pursue testing. It is essential that practitioners continue to advocate for clinical genetic testing guidelines that better reflect the state of the research on genetics in ASD.

## Data Availability

Genetic and phenotypic data were obtained through SFARI Base. Approved researchers can obtain the SSC population dataset described in this study by applying at https://base.sfari.org.

## Acknowledgments

This work was supported by grants from the Autism Science Foundation (ASP-16-001, S.L.B., E.R., S.J.S.) and the NIMH (RMH111813A to E.R; U01 MH111662, S.J.S.), as well as the NIMH Intramural Research Program (1ZICMH002961, A.T).

Somer Bishop receives royalties from Western Psychological Services (WPS) for sales of the Autism Diagnostic Observation Schedule, 2nd Edition (ADOS-2) manual and protocols. All other authors declare no conflict of interest.

We are grateful to all of the families at the participating Simons Simplex Collection (SSC) sites, as well as the principal investigators (A. Beaudet, R. Bernier, J. Constantino, E. Cook, E. Fombonne, D. Geschwind, R. Goin-Kochel, E. Hanson, D. Grice, A. Klin, D. Ledbetter, C. Lord, C. Martin, D. Martin, R. Maxim, J. Miles, O. Ousley, K. Pelphrey, B. Peterson, J. Piggot, C. Saulnier, M. State, W. Stone, J. Sutcliffe, C. Walsh, Z. Warren, E. Wijsman). We appreciate obtaining access to genetic and phenotypic data on SFARI Base. Approved researchers can obtain the SSC population dataset described in this study by applying at https://base.sfari.org.

The views expressed in this article do not necessarily represent the views of the National Institutes of Health, the Department of Health and Human Services, or the United States Government.

