## Supplementary material for "Prevalence of Returnable Genetic Results Based on Recognizable Phenotypes among Children with Autism Spectrum Disorder": Table S1

| Sex | ID | Walking | Seizures | Any_Count | Yes_Count | No_Count | Total | Returnable | Percent_Returnable | Percent_of_ASD_cases | Percent_of_returnable_cases | Number_needed_to_return |
|---|---|---|---|---|---|---|---|---|---|---|---|---|
| Any | Any | Any | Any | 4 | 0 | 0 | 2218 | 234 | 10.60% | 100.00% | 100.00% | 9.5 |
| Any | Any | Any | No | 3 | 0 | 1 | 2102 | 200 | 9.50% | 94.80% | 85.50% | 10.5 |
| Any | Any | Any | Yes | 3 | 1 | 0 | 116 | 34 | 29.30% | 5.20% | 14.50% | 3.4 |
| Any | Any | No | Any | 3 | 0 | 1 | 1972 | 157 | 8.00% | 88.90% | 67.10% | 12.6 |
| Any | Any | No | No | 2 | 0 | 2 | 1881 | 138 | 7.30% | 84.80% | 59.00% | 13.6 |
| Any | Any | No | Yes | 2 | 1 | 1 | 91 | 19 | 20.90% | 4.10% | 8.10% | 4.8 |
| Any | Any | Yes | Any | 3 | 1 | 0 | 246 | 77 | 31.30% | 11.10% | 32.90% | 3.2 |
| Any | Any | Yes | No | 2 | 1 | 1 | 221 | 62 | 28.10% | 10.00% | 26.50% | 3.6 |
| Any | Any | Yes | Yes | 2 | 2 | 0 | 25 | 15 | 60.00% | 1.10% | 6.40% | 1.7 |
| Any | Yes | Any | Any | 3 | 1 | 0 | 664 | 108 | 16.30% | 29.90% | 46.20% | 6.1 |
| Any | Yes | Any | No | 2 | 1 | 1 | 599 | 84 | 14.00% | 27.00% | 35.90% | 7.1 |
| Any | Yes | Any | Yes | 2 | 2 | 0 | 65 | 24 | 36.90% | 2.90% | 10.30% | 2.7 |
| Any | Yes | No | Any | 2 | 1 | 1 | 557 | 61 | 11.00% | 25.10% | 26.10% | 9.1 |
| Any | Yes | No | No | 1 | 1 | 2 | 513 | 50 | 9.70% | 23.10% | 21.40% | 10.3 |
| Any | Yes | No | Yes | 1 | 2 | 1 | 44 | 11 | 25.00% | 2.00% | 4.70% | 4.0 |
| Any | Yes | Yes | Any | 2 | 2 | 0 | 107 | 47 | 43.90% | 4.80% | 20.10% | 2.3 |
| Any | Yes | Yes | Yes | 1 | 3 | 0 | 21 | 13 | 61.90% | 0.90% | 5.60% | 1.6 |
| Any | Yes | Yes | No | 1 | 2 | 1 | 86 | 34 | 39.50% | 3.90% | 14.50% | 2.5 |
| Any | No | Any | Any | 3 | 0 | 1 | 1554 | 126 | 8.10% | 70.10% | 53.80% | 12.3 |
| Any | No | Any | No | 2 | 0 | 2 | 1503 | 116 | 7.70% | 67.80% | 49.60% | 13.0 |
| Any | No | Any | Yes | 2 | 1 | 1 | 51 | 10 | 19.60% | 2.30% | 4.30% | 5.1 |
| Any | No | No | Any | 2 | 0 | 2 | 1415 | 96 | 6.80% | 63.80% | 41.00% | 14.7 |
| Any | No | No | No | 1 | 0 | 3 | 1368 | 88 | 6.40% | 61.70% | 37.60% | 15.5 |
| Any | No | No | Yes | 1 | 1 | 2 | 47 | 8 | 17.00% | 2.10% | 3.40% | 5.9 |
| Any | No | Yes | Any | 2 | 1 | 1 | 139 | 30 | 21.60% | 6.30% | 12.80% | 4.6 |
| Any | No | Yes | No | 1 | 1 | 2 | 135 | 28 | 20.70% | 6.10% | 12.00% | 4.8 |
| Any | No | Yes | Yes | 1 | 2 | 1 | 4 | 2 | 50.00% | 0.20% | 0.90% | 2.0 |
| Male | Any | Any | Any | 3 | 0 | 0 | 1927 | 177 | 9.20% | 86.90% | 75.60% | 10.9 |
| Male | Any | Any | No | 2 | 0 | 1 | 1832 | 153 | 8.40% | 82.60% | 65.40% | 12.0 |
| Male | Any | Any | Yes | 2 | 1 | 0 | 95 | 24 | 25.30% | 4.30% | 10.30% | 4.0 |
| Male | Any | No | Any | 2 | 0 | 1 | 1731 | 119 | 6.90% | 78.00% | 50.90% | 14.5 |
| Male | Any | No | No | 1 | 0 | 2 | 1654 | 107 | 6.50% | 74.60% | 45.70% | 15.5 |
| Male | Any | No | Yes | 1 | 1 | 1 | 77 | 12 | 15.60% | 3.50% | 5.10% | 6.4 |
| Male | Any | Yes | Any | 2 | 1 | 0 | 196 | 58 | 29.60% | 8.80% | 24.80% | 3.4 |
| Male | Any | Yes | No | 1 | 1 | 1 | 178 | 46 | 25.80% | 8.00% | 19.70% | 3.9 |
| Male | Any | Yes | Yes | 1 | 2 | 0 | 18 | 12 | 66.70% | 0.80% | 5.10% | 1.5 |
| Male | Yes | Any | Any | 2 | 1 | 0 | 544 | 72 | 13.20% | 24.50% | 30.80% | 7.6 |
| Male | Yes | Any | No | 1 | 1 | 1 | 496 | 57 | 11.50% | 22.40% | 24.40% | 8.7 |
| Male | Yes | Any | Yes | 1 | 2 | 0 | 48 | 15 | 31.20% | 2.20% | 6.40% | 3.2 |
| Male | Yes | No | Any | 1 | 1 | 1 | 467 | 40 | 8.60% | 21.10% | 17.10% | 11.7 |
| Male | Yes | No | No | 0 | 1 | 2 | 434 | 35 | 8.10% | 19.60% | 15.00% | 12.4 |

| | | | | | | | | | | | | |
|---|---|---|---|---|---|---|---|---|---|---|---|---|
| Male | Yes | No | Yes | 0 | 2 | 1 | 33 | 5 | 15.20% | 1.50% | 2.10% | 6.6 |
| Male | Yes | Yes | Any | 1 | 2 | 0 | 77 | 32 | 41.60% | 3.50% | 13.70% | 2.4 |
| Male | Yes | Yes | Yes | 0 | 3 | 0 | 15 | 10 | 66.70% | 0.70% | 4.30% | 1.5 |
| Male | Yes | Yes | No | 0 | 2 | 1 | 62 | 22 | 35.50% | 2.80% | 9.40% | 2.8 |
| Male | No | Any | Any | 2 | 0 | 1 | 1383 | 105 | 7.60% | 62.40% | 44.90% | 13.2 |
| Male | No | Any | No | 1 | 0 | 2 | 1336 | 96 | 7.20% | 60.20% | 41.00% | 13.9 |
| Male | No | Any | Yes | 1 | 1 | 1 | 47 | 9 | 19.10% | 2.10% | 3.80% | 5.2 |
| Male | No | No | Any | 1 | 0 | 2 | 1264 | 79 | 6.20% | 57.00% | 33.80% | 16.0 |
| Male | No | No | No | 0 | 0 | 3 | 1220 | 72 | 5.90% | 55.00% | 30.80% | 16.9 |
| Male | No | No | Yes | 0 | 1 | 2 | 44 | 7 | 15.90% | 2.00% | 3.00% | 6.3 |
| Male | No | Yes | Any | 1 | 1 | 1 | 119 | 26 | 21.80% | 5.40% | 11.10% | 4.6 |
| Male | No | Yes | No | 0 | 1 | 2 | 116 | 24 | 20.70% | 5.20% | 10.30% | 4.8 |
| Male | No | Yes | Yes | 0 | 2 | 1 | 3 | 2 | 66.70% | 0.10% | 0.90% | 1.5 |
| Female | Any | Any | Any | 3 | 0 | 0 | 291 | 57 | 19.60% | 13.10% | 24.40% | 5.1 |
| Female | Any | Any | No | 2 | 0 | 1 | 270 | 47 | 17.40% | 12.20% | 20.10% | 5.7 |
| Female | Any | Any | Yes | 2 | 1 | 0 | 21 | 10 | 47.60% | 0.90% | 4.30% | 2.1 |
| Female | Any | No | Any | 2 | 0 | 1 | 241 | 38 | 15.80% | 10.90% | 16.20% | 6.3 |
| Female | Any | No | No | 1 | 0 | 2 | 227 | 31 | 13.70% | 10.20% | 13.20% | 7.3 |
| Female | Any | No | Yes | 1 | 1 | 1 | 14 | 7 | 50.00% | 0.60% | 3.00% | 2.0 |
| Female | Any | Yes | Any | 2 | 1 | 0 | 50 | 19 | 38.00% | 2.30% | 8.10% | 2.6 |
| Female | Any | Yes | No | 1 | 1 | 1 | 43 | 16 | 37.20% | 1.90% | 6.80% | 2.7 |
| Female | Any | Yes | Yes | 1 | 2 | 0 | 7 | 3 | 42.90% | 0.30% | 1.30% | 2.3 |
| Female | No | Any | Any | 2 | 0 | 1 | 171 | 21 | 12.30% | 7.70% | 9.00% | 8.1 |
| Female | No | Any | No | 1 | 0 | 2 | 167 | 20 | 12.00% | 7.50% | 8.50% | 8.4 |
| Female | No | Any | Yes | 1 | 1 | 1 | 4 | 1 | 25.00% | 0.20% | 0.40% | 4.0 |
| Female | No | No | Any | 1 | 0 | 2 | 151 | 17 | 11.30% | 6.80% | 7.30% | 8.9 |
| Female | No | No | No | 0 | 0 | 3 | 148 | 16 | 10.80% | 6.70% | 6.80% | 9.3 |
| Female | No | No | Yes | 0 | 1 | 2 | 3 | 1 | 33.30% | 0.10% | 0.40% | 3.0 |
| Female | No | Yes | Any | 1 | 1 | 1 | 20 | 4 | 20.00% | 0.90% | 1.70% | 5.0 |
| Female | No | Yes | No | 0 | 1 | 2 | 19 | 4 | 21.10% | 0.90% | 1.70% | 4.8 |
| Female | No | Yes | Yes | 0 | 2 | 1 | 1 | 0 | 0.00% | 0.00% | 0.00% | NA |
| Female | Yes | Any | Any | 2 | 1 | 0 | 120 | 36 | 30.00% | 5.40% | 15.40% | 3.3 |
| Female | Yes | Any | No | 1 | 1 | 1 | 103 | 27 | 26.20% | 4.60% | 11.50% | 3.8 |
| Female | Yes | Any | Yes | 1 | 2 | 0 | 17 | 9 | 52.90% | 0.80% | 3.80% | 1.9 |
| Female | Yes | Yes | Any | 1 | 2 | 0 | 30 | 15 | 50.00% | 1.40% | 6.40% | 2.0 |
| Female | Yes | Yes | No | 0 | 2 | 1 | 24 | 12 | 50.00% | 1.10% | 5.10% | 2.0 |
| Female | Yes | Yes | Yes | 0 | 3 | 0 | 6 | 3 | 50.00% | 0.30% | 1.30% | 2.0 |
| Female | Yes | No | Any | 1 | 1 | 1 | 90 | 21 | 23.30% | 4.10% | 9.00% | 4.3 |
| Female | Yes | No | No | 0 | 1 | 2 | 79 | 15 | 19.00% | 3.60% | 6.40% | 5.3 |
| Female | Yes | No | Yes | 0 | 2 | 1 | 11 | 6 | 54.50% | 0.50% | 2.60% | 1.8 |
